# Gender and Belief about Menstruation and Academic Performance

**DOI:** 10.1101/2023.11.26.23299031

**Authors:** Destiny Odah

**Affiliations:** Department of Health Sciences and Social Work, Western Illinois University, Macomb, IL, U.S.A

## Abstract

Menstruation is a natural process for women that shows a healthy female reproductive system, yet women are stigmatized during this experience. A woman’s physical, mental, and social well-being are significantly impacted by menstruation. Thus, menstrual disorders are widespread and are a major social problem. There have been variations in people’s perceptions of menstruation and gaps in research comparing gender and beliefs about menstruation and its impact on academic performance. This study investigates gender and beliefs about menstruation and how it affects academic performance and social life using a self-administered online questionnaire that inquires about general knowledge of menstruation, participants’ beliefs and experiences with menstruation, participants’ academic performance and demographic questions. Participants responded using a 5-point Likert scale, showing that 40.44% of the respondents’ knowledge and understanding of menstruation have positively influenced over the years. Furthermore, the results from the chi-square analysis revealed an association between men’s and women’s perceptions of menstruation. Interestingly, women stated that menstruation has no effect on their academic performances, but on the other hand, men were indecisive about its impact on women’s intellectual tasks. Also, 28.72% of respondents reported that they find studying and excelling during menstruation uneasy. The result shows that women can excel at any task, even during menstruation. Knowledge of menstruation is essential in furthering a better understanding of women’s health, demystifying menstruation myths, and advancing the social well-being of this gender.

## Introduction

Menstruation is a naturally occurring physiological phenomenon in women. It is controlled in a healthy woman by cyclical changes in estrogen and progesterone levels, which happen periodically throughout a cycle that lasts around one month (Farage *et al*., 2009). Due to considerable hormonal changes, many changes in a woman’s body occur throughout the menstrual cycle, from the first day of menstruation to the day before the next menstrual period. The effects of menstruation on a woman’s physical, emotional, and social health are profound. “A degree of total physical, mental, and social well-being and not only the absence of illness or dysfunction about the monthly cycle” defines menstrual health (Hennegan *et al*., 2021). Therefore, menstrual symptoms are among women’s most prevalent issues (Farage *et al*., 2009)—several variables relating to physical changes are viewed as disturbing and anguishing.

Premenstrual syndrome, or PMS as it is more generally known, remains the central area of interest in menstrual cycle research. Greene and Dalton (1953) introduced premenstrual syndrome to describe painful physical and psychological changes before and during menstruation. Premenstrual symptoms occur before menstruation, particularly during the luteal phase, and are associated with various cognitive, behavioral, and psychological changes (Vahia, 2013). Symptoms of premenstrual syndrome include irritability, anxiety, disorientation, depression, rage outburst, or social withdrawal. Physiological symptoms are abdominal bloating, breast tenderness, headache, or swelling of extremities. These symptoms occurring during the five days preceding menstruation and occurring in at least three consecutive menstrual cycles signify PMS (Mortola *et al*., 1989). These symptoms generally subside four days after the start of menstruation.

Premenstrual dysphoric disorder (PMDD) is a severe type of premenstrual syndrome (PMS) that affects 3-8% of women and causes significant psychological symptoms (Dillaz & Aksan, 2021). PMDD is described as having one or more of the following symptoms, according to the Diagnostic and Statistical Manual of Mental Disorders, Fifth Edition criteria: mood changes or sudden sadness or increased sensitivity to rejection, anger or irritability, sense of hopelessness or depression, self-critical thoughts, and anxiety and feeling on edge (Hantsoo & Epperson, 2015). Additional signs include a loss of interest, difficulty concentrating, exhaustion, changes in eating and sleep patterns, a sense of being out of control, and physical symptoms. These physical signs include breast soreness, swelling, joint or muscle pain, and weight gain. These symptoms negatively impact social and professional life and are attributed to other material or psychiatric illnesses or drugs (Shehadeh & Hamdan-Mansour, 2018).

Dysmenorrhea (primary and secondary), the term for menstrual aches and pains, is another typical menstrual condition. Secondary dysmenorrhea, in contrast to initial dysmenorrhea, is linked to pelvic pathology. A study by Alshahrani in 2020 reported that dysmenorrhea affects 15.8 to 89.5% of women, with 20% reporting severe symptoms. However, secondary dysmenorrhea can occur due to gynecologic disorders such as endometriosis, adenomyosis, or uterine fibroid. Primary dysmenorrhea is reoccurring lower abdominal and pelvic pain during menstruation, affecting 45 to 95% of women of reproductive age. Although finding no correlation between dysmenorrhea and physical activity or skipping breakfast, a study (Faramarzi & Salmalian, 2014) found that women with higher levels of stress and coffee consumption had a higher risk of the condition. According to (Hu *et al*.,2020), women who skip breakfast, have short sleep durations, and go to bed late are likelier to experience dysmenorrhea than those who engage in physical activity or consume caffeine.

There have been variations and gaps in people’s perceptions of menstruation. In a study carried out in Canada on women’s perception of menstruation and PMS, several women’s responses showed a positive and negative attitude toward the meaning of menstruation (Lee, 2002). Bhartiya, (2013) showed that gender, religion, and culture frequently establish menstrual customs and beliefs. Menstruation is socially unacceptable or stigmatized in many parts of the world, particularly in Asia and Africa’s low-and middle-income nations (Hennegan *et al*.,2019). The concept of sickness as an undesirable condition in medical anthropological work adversely affects how women view their menstrual cycle (Hahn, 1995). Hahn (1995) also mentioned that a person’s sense of self is thought to be threatened by sickness centered on their experience of a disordered state. As a result, a person devalues an undesirable situation based on specific characteristics they have learned to identify with health or well-being. According to Lee (2002), menstruation might be considered a negative or unpleasant condition (called PMS) or have a more positive implication. Women’s behavior and work capacity have traditionally been thought to be influenced by menstruation and the menstrual cycle. Also connected to lower levels of body acceptance are negative attitudes concerning menstruation (Schooler, Ward, Meriwether, & Caruthers, 2005). The authors utilized computational modeling of structural equations on a sample of 199 undergraduate women to discover that women less secure with their bodies expressed reduced positive perceptions of menstruation and were less forceful regarding sexual decision-making (Schooler *et al*., 2005).

Men’s perceptions of menstruation were explored in Fishman’s 2014 thesis paper. It was found that most participants said that knowing about menstruation in their early years left them with the strong impression that it is not a subject that ought to be addressed. Many participants reported that their opinions about menstruation as adults remained shaped by their menstrual experiences as children, maybe because they lacked the desire to reevaluate harmful beliefs. According to Hamdan-Mansour *et al*. (2012), university students are at a stage in life where they lack the psychological and social skills necessary to manage daily life stressors. Despite lacking resources and being plagued by psychological and social expectations, they endeavor for more outstanding educational accomplishments to obtain better employment and meet their actualization goals (Hamdan-Mansour *et al*., 2012; Neumann *et al*., 2009). The impact of menstruation on academic performance has been researched for the past century. According to a recent comprehensive review, dysmenorrhea is associated with academic impairments such as absenteeism, less engagement in class activities, lack of concentration, and poor academic performance (Munro *et al*., 2021). Although studies have reported dysmenorrhea as associated with intellectual impairments, no study has compared gender beliefs about menstruation and its impacts on academic performance and social activities.

This study aims to establish the beliefs and perceptions of menstruation and its effects on female academic performance and social activities.

## Material & Methods

A self-administered online survey was used —the questionnaire comprised four sections. The first section inquired about general knowledge of menstruation; the second section asked about participants’ beliefs and experiences with menstruation; the third investigated participants’ academic performance; the last section was on socio-demographic questions. Participants responded using a 5-point Likert scale response. The Western Illinois University Institutional Review Board (IRB) approved this study. An email solicitation was sent to Western Illinois University students with the survey links. Western Illinois University students aged 18 and above interested in proceeding to the questionnaire had to provide online informed consent. They were informed of their right to withdraw at any time, and no risk was involved in participating. The survey was sent to students a month before the end of the spring 2023 semester, of which 280 students responded. No identification information was gathered.

Before analysis, the dataset was checked for missing data and then analyzed using the statistical software SPSS. Also, the chi-square statistical test was used to examine the correlation between gender’s perception of menstruation.

## Result and Discussion

### Demographic characteristics

A total of 264 students completed this section (Table 1), of which all declared their age, 261 revealed their race/ ethnicity, and 262 revealed their gender identity. The highest number of participants in this study falls in the age group 18-22 (n=134, 50.76%), followed by the age group 23-27 (n=58, 21.97%), compared to participants older than 42 (n=26, 9.85%), showing that participants are college students. Most participants are white (n=191, 73.18%); however, the participants cut across different ethnicities. Women are more prevalent (n=188, 71.76%) as the majority of respondents compared to the other genders, which can be expected. Also, the participants specialize in several majors at the university.

**Table 1.**
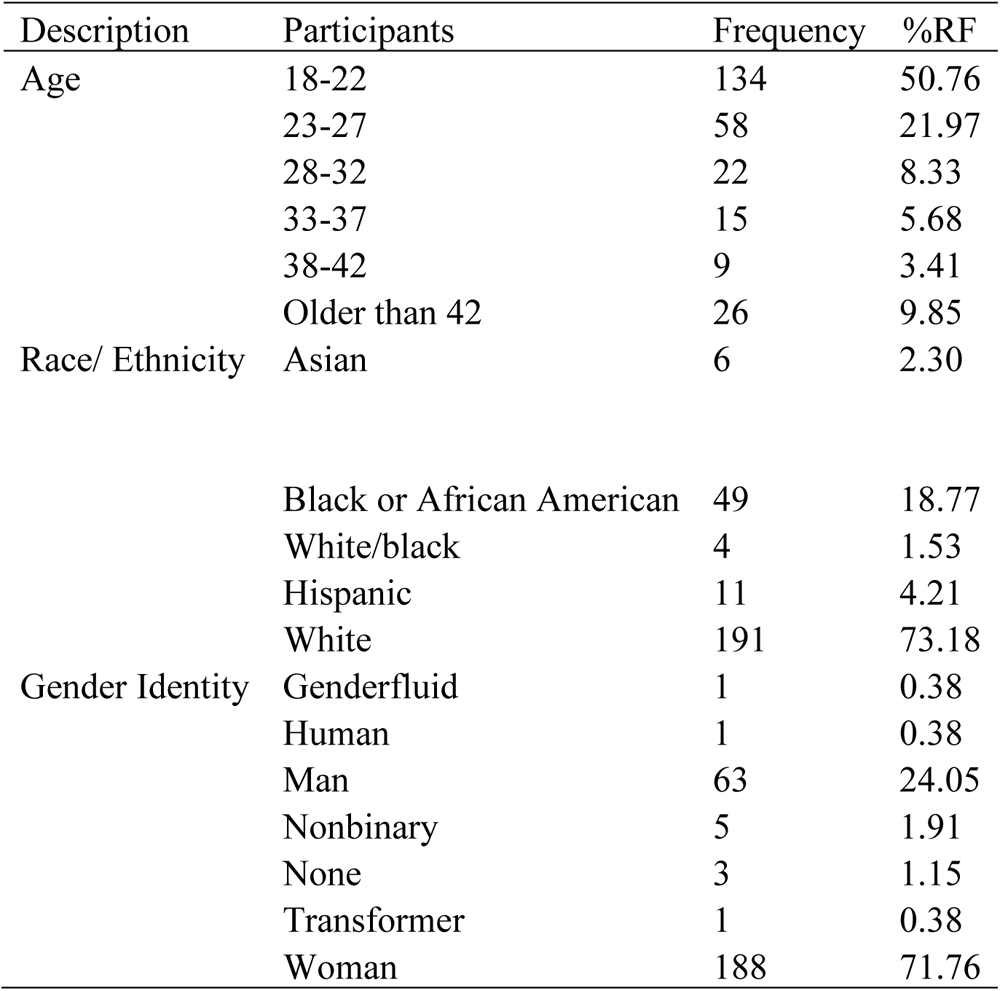
Demographic information of participants (N=264)

### Knowledge of menstruation

This section aims to examine participants’ general knowledge of menstruation. A total of 143 (54.6%) of the 262 participants reported that they were between the ages 6-10 when they first heard about menstruation (Fig 1), which suggests that they have a good knowledge of menstruation at an early age. Furthermore, the source of participant’s early education on menstruation was further explored, and the result from the survey showed that most participants (37.5%) acquired menstruation information from family members and siblings (Fig 2), suggesting effective communication of menstruation among parents and children. Also, this study examines participants’ knowledge of menstruation over time. The survey showed that the understanding of most participants (40.44%, n= 110) had been positively influenced over time (Table 2). Although this study did not go into detail about the positive change in knowledge over time, the result suggests that the mindset of participants has been positively influenced as they grow.

**Figure 1.**
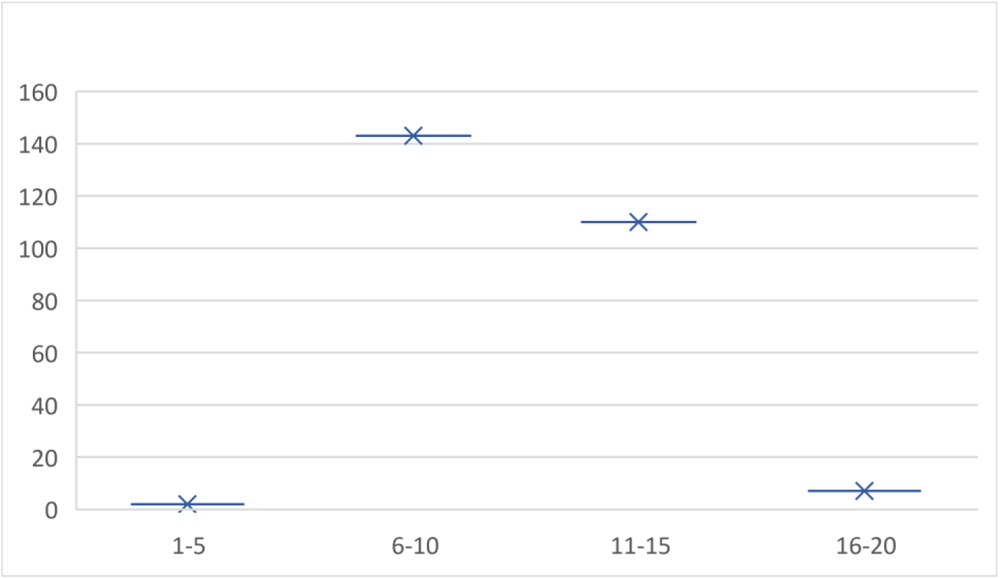
The age range at which participants first heard about menstruation.

**Figure 2.**
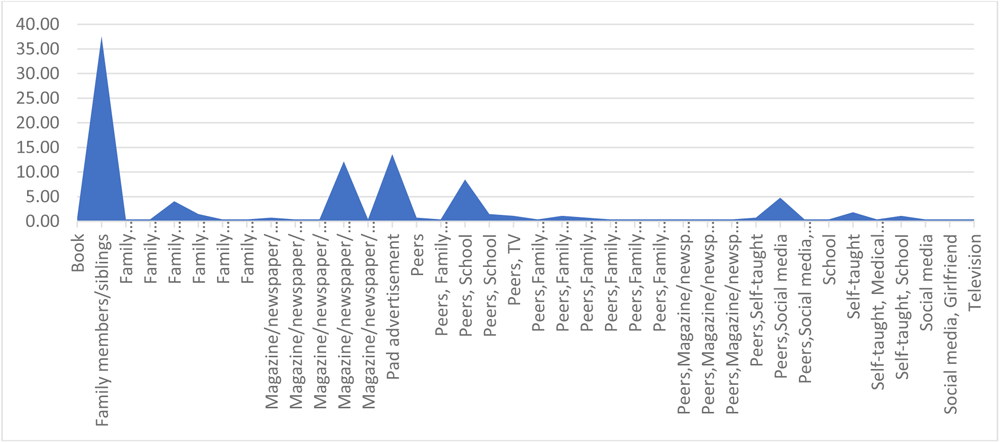
Participants’ source of menstruation information.

**Table 2.** Change in participants knowledge about menstruation over time (N=272).

| Change in menstruation Knowledge | Frequency | %RF |
| --- | --- | --- |
| Negative | 11 | 4.04 |
| Neutral | 69 | 25.37 |
| Positive | 110 | 40.44 |
| Strongly negative | 4 | 1.47 |
| Strongly positive | 78 | 28.68 |

### Beliefs and Perceptions

In this section, the perception of men and women were examined singly to understand each gender’s belief of menstruation properly. A total of 63 men (Table 3) participated in the survey, while 190 women participated (Table 4). Although most men participants (n= 31, 49.21%) agree that tiredness seen in women during menstruation could be associated with dysmenorrhea, a significant number of men participants do not agree that women’s fatigue could be related to menstruation (n= 30, 47.62%). On the other hand, an increased number of women participants (47.37, n=90) agreed that menstruation complications could be related to tiredness seen in women during menstruation. Interestingly, 71 women participants (37.37%) strongly agreed, suggesting that men might be unaware of the exhaustion associated with women’s menstruation. Furthermore, the study examines the participants’ perception of the effect of menstruation on social activities. The feedback revealed that 89 (46.84%) and 75 (39.47%) woman participants agreed and strongly agreed to turn down social plans due to menstruation, which is related to men participants’ responses. Notably, most women participants reported that menstruation does not affect their intellectual tasks (strongly disagree = 60, 31.58%), while most men were indecisive (neutral = 23, 36.51%). The statistical package for the social sciences (SPSS) was used to carry out a chi-square to determine whether there is a relationship between men’s and women’s perceptions. The result showed that there is an association between the beliefs of the two genders (p-value < 0.05) (Table 5).

**Table 3.**
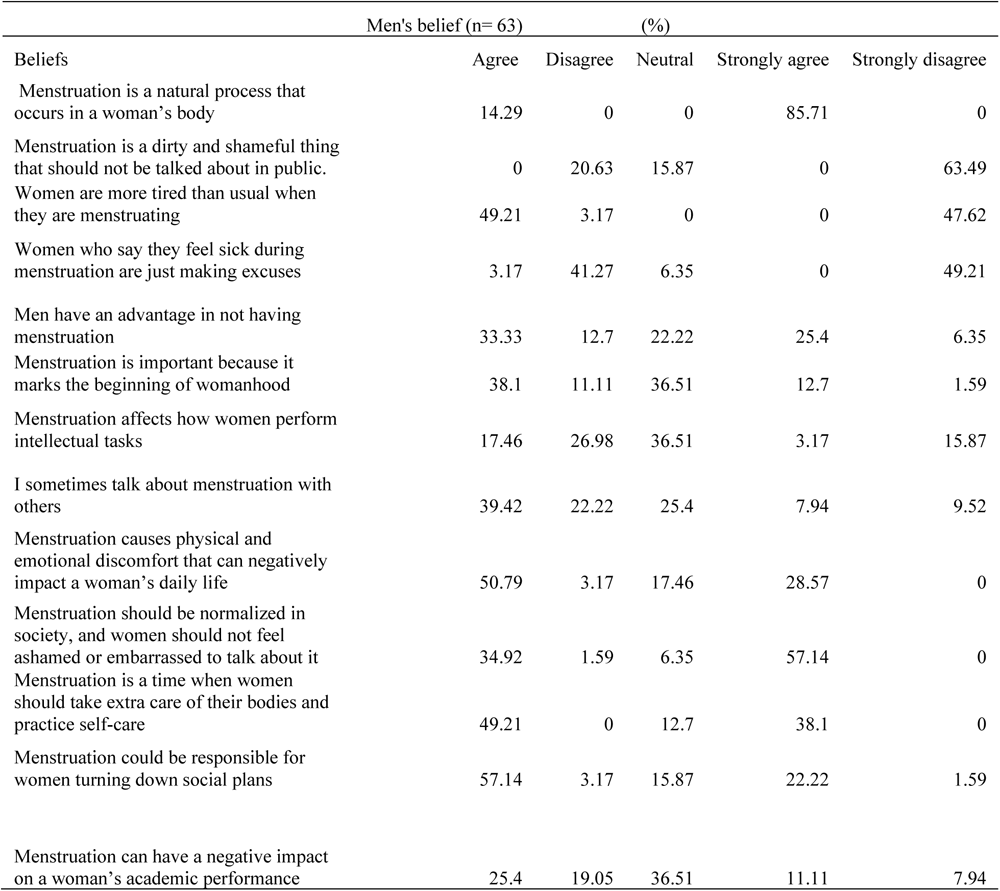
Men’s belief and perception about menstruation.

| Beliefs | Men's belief (n= 63) |  |  |  |  |
| --- | --- | --- | --- | --- | --- |
|  | (%) |  |  |  |  |
|  | Agree | Disagree | Neutral | Strongly agree | Strongly disagree |
| Menstruation is a natural process that occurs in a woman's body | 14.29 | 0 | 0 | 85.71 | 0 |
| Menstruation is a dirty and shameful thing that should not be talked about in public. | 0 | 20.63 | 15.87 | 0 | 63.49 |
| Women are more tired than usual when they are menstruating | 49.21 | 3.17 | 0 | 0 | 47.62 |
| Women who say they feel sick during menstruation are just making excuses | 3.17 | 41.27 | 6.35 | 0 | 49.21 |
| Men have an advantage in not having menstruation | 33.33 | 12.7 | 22.22 | 25.4 | 6.35 |
| Menstruation is important because it marks the beginning of womanhood | 38.1 | 11.11 | 36.51 | 12.7 | 1.59 |
| Menstruation affects how women perform intellectual tasks | 17.46 | 26.98 | 36.51 | 3.17 | 15.87 |
| I sometimes talk about menstruation with others | 39.42 | 22.22 | 25.4 | 7.94 | 9.52 |
| Menstruation causes physical and emotional discomfort that can negatively impact a woman's daily life | 50.79 | 3.17 | 17.46 | 28.57 | 0 |
| Menstruation should be normalized in society, and women should not feel ashamed or embarrassed to talk about it | 34.92 | 1.59 | 6.35 | 57.14 | 0 |
| Menstruation is a time when women should take extra care of their bodies and practice self-care | 49.21 | 0 | 12.7 | 38.1 | 0 |
| Menstruation could be responsible for women turning down social plans | 57.14 | 3.17 | 15.87 | 22.22 | 1.59 |
| Menstruation can have a negative impact on a woman's academic performance | 25.4 | 19.05 | 36.51 | 11.11 | 7.94 |

**Table 4.**
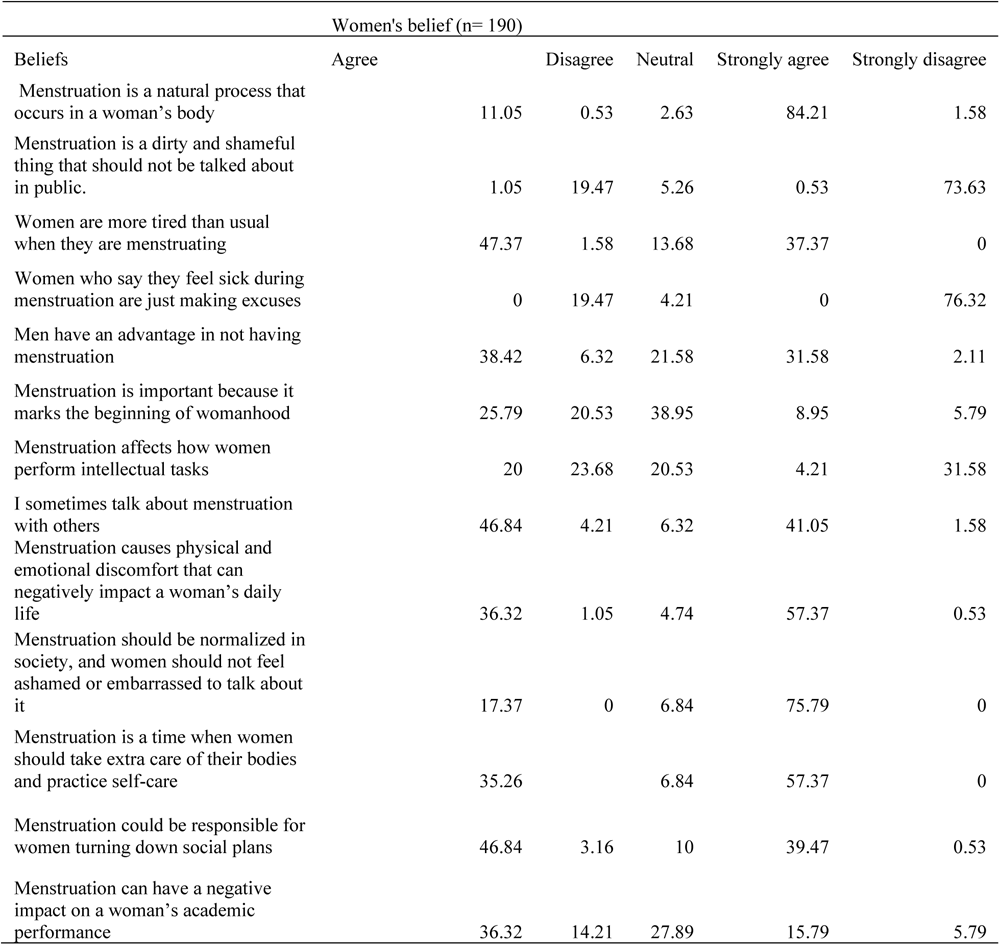
Women’s belief and perception about menstruation.

**Table 5.** Relationship between men and female perception about menstruation (95% CI)

| **CHI-SQUARE** |  |  |
| --- | --- | --- |
| **Belief and Perception** | **df** | **Pvalue** |
| Menstruation is a natural process that occurs in a woman's body | 16 | 2.20E-12 |
| Menstruation is a dirty and shameful thing that should not be talked about in public. | 12 | 2.20E-16 |
| Women are more tired than usual when they are menstruating | 12 | 2.20E-16 |
| Women who say they feel sick during menstruation are just making excuses | 10 | 2.20E-16 |
| Men have an advantage in not having menstruation | 20 | 2.20E-16 |
| Menstruation is important because it marks the beginning of womanhood | 20 | 2.20E-16 |
| Menstruation affects how women perform intellectual tasks | 20 | 2.20E-16 |
| I sometimes talk about menstruation with others | 20 | 7.02E-14 |
| Menstruation causes physical and emotional discomfort that can negatively impact a woman's daily life | 16 | 2.20E-16 |
| Menstruation should be normalized in society, and women should not feel ashamed or embarrassed to talk about it | 8 | 2.20E-16 |
| Menstruation is a time when women should take extra care of their bodies and practice self-care | 9 | 2.20E-16 |
| Menstruation could be responsible for women turning down social plans | 20 | 7.02E-14 |
| Menstruation can have a negative impact on a woman's academic performance | 20 | 2.20E-16 |

**Table 6.** Participant grade point average.

|  | Range | Frequency | %RF |
| --- | --- | --- | --- |
| Participant GPA | 1-1.9 | 1 | 0.51 |
|  | 2-2.9 | 16 | 8.21 |
|  | 3-4 | 174 | 89.23 |

### Academic performance

The academic performance of students before and during menstruation was further explored to examine any association. First, students’ academic performance before menstruation was examined, and the highest population (36.92%, n=72) agreed that studying and excelling at any academic task before menstruation was easy (Figure 3). Next, the study investigated how easy studying and excelling at any academic performance was during menstruation. The result showed that 28.72% (n=56) of participants reported feeling uneasy about studying and excelling during menstruation—also, 28.72% (n=56) were indecisive about how they perform at their academic task during menstruation, suggesting that it might not be very easy for them during their periods (Figure 3). Furthermore, participants’ satisfaction with the results of exams taken during menstruation was examined. The results showed that the highest number of participants was uncertain (35.9%, n= 70), suggesting their results might not be as good as those taken without menstruation. On the other hand, 34.87% (n=68) reported that they are satisfied regardless of their menstruation (Figure 3). To further clarify, the survey explored if students’ academic performance goes as expected during menstruation. The results showed that the most significant number of participants (45.13%, n=88) reported that menstruation does not affect their academic performance (Figure 3). Interestingly, 89.23% (n=174) of the participant’s GPA falls in the range of 3-4 (Table 5). Also, a significant number of participants (45.54%, n=89) reported that they don’t pile up their school workload till after menstruation, suggesting that students can perform well in their academic tasks during menstruation even though they find it uneasy. Lastly, the degree of absenteeism due to dysmenorrhea was examined among participants (Figure 4). The results showed that many participants (59%, n=111) were never absent from school due to menstruation complications, which further validated that students can resist the intricacies of menstruation. Notably, other participants reported their absenteeism, suggesting that complications might differ among women.

**Figure 3.**
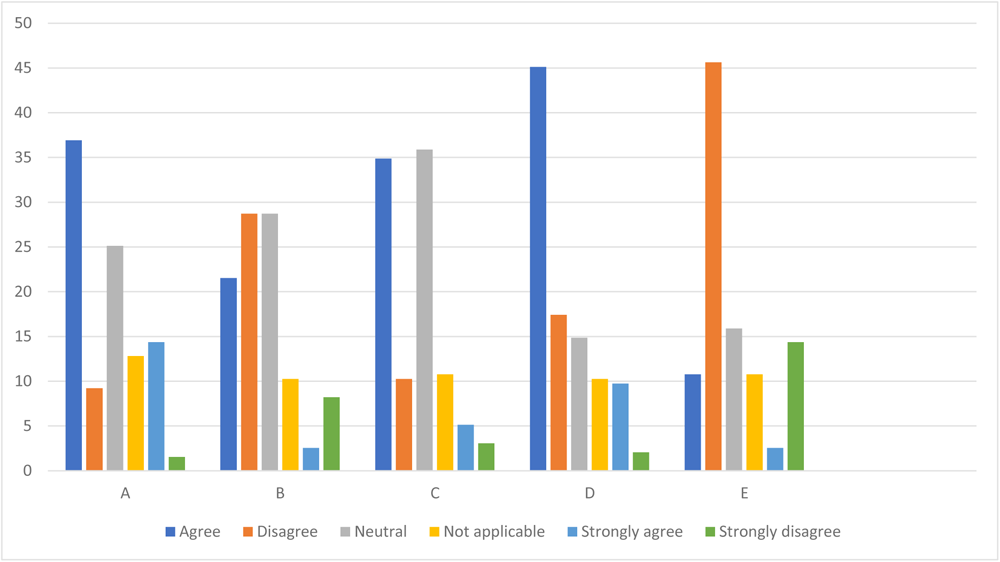
Participants’ academic performance. A - I find studying and excelling at any academic task “before” menstruation easy; B - I find studying and excelling at any academic task “during” menstruation easy; C - I am satisfied with the results of exams taken during menstruation; D - During menstruation, my academic performance goes as expected; E - After menstruation, my academic workload piles up so high that I cannot overcome them.

**Figure 4.**
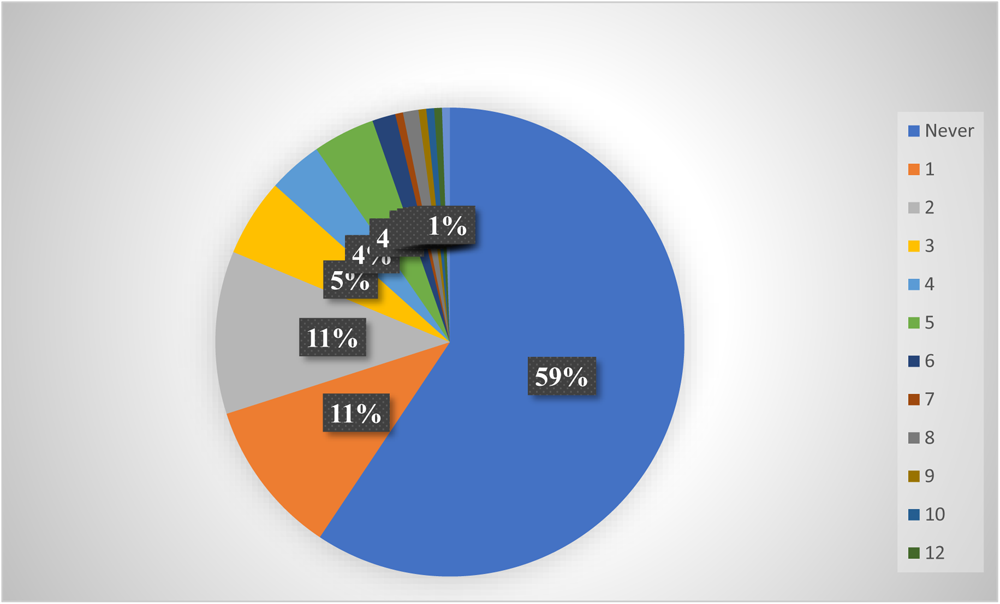
Participants absenteeism due to dysmenorrhea.

## Conclusion

Through a self-administered online survey, this study revealed that men are indecisive about menstruation complications on females’ academic performance. Also, women reported that menstruation does not affect their academic performance or ability to perform intellectual tasks despite their struggles with fatigue during menstruation. However, menstruation could be associated with women turning social plans down. Future studies should examine menstruation knowledge of men and disembark information to fill the knowledge gap. Also, previous studies have reported that menstruation affects women’s academic performance; however, this study showed that menstruation does not affect women’s academic performance but agrees that it does affect their social life. Thus, future studies should further examine this to resolve the controversy.

## Data Availability

All data produced in the present study are available upon reasonable request to the authors

## Acknowledgments

I would love to thank Dr. Lora Wallace for reviewing and assisting with this study.

